# Evaluating the impact of RSV immunisation strategies on antibiotic use in England

**DOI:** 10.1101/2021.11.08.21266072

**Authors:** Katherine E. Atkins, David Hodgson, Mark Jit, Nicholas G. Davies

**Affiliations:** London School of Hygiene and Tropical Medicine, London, UK; University of Edinburgh, Scotland, UK

**Author notes:** Corresponding author. Address for correspondence: Katherine E. Atkins, Centre for Global Health, Usher Institute, MacKenzie House, 30 West Richmond Street, University of Edinburgh, EH8 9DX, United Kingdom.

## Abstract

With a sizable fraction of primary care antibiotics prescribing attributable to RSV, the promising suite of prophylactic pharmaceuticals against could reduce the need for antibiotics in addition to controlling respiratory disease. To assess the potential impact of RSV vaccines on the reduction in primary care antibiotic prescribing in England, we integrate results from a dynamic transmission model of RSV and a statistical attribution framework. Under base case assumptions, targeting children aged 5-14 years reduces antibiotic prescribing by 10.9 (8.0-14.2) antibiotic courses per 10,000 person years. This reduction in antibiotic use would gain 128 DALYs and avert 51,000 GBP associated with infections caused by drug resistant bacteria. Seasonally administering monoclonal antibodies (mAbs) to high risk infants under 6 months is the most efficient strategy (reducing per person year antibiotic prescribing by 2.6 (1.9-3.3) antibiotic courses per 1,000 mAb courses).

## Introduction

Vaccines against viral pathogens have been suggested as a novel means to reduce antibiotic resistance (1). By reducing the number of viral infections, fewer antibiotics would be used either inappropriately against viral disease, as a precautionary measure for non-specific symptoms, or to treat bacterial co-infections (2,3). Consequently, this reduction in antibiotic use would exert less selection for resistance on highly prevalent commensal bacteria that can lead to invasive disease.

Respiratory syncytial virus (RSV) is a major cause of acute lower respiratory tract infections in young children globally (4). RSV can cause mild upper respiratory tract symptoms across all age groups and these infections can often result in primary care visits (5). Data from high income countries suggest that antibiotic prescribing is common amongst primary care visits attributable to RSV infection, and consequently, reducing RSV infections may be beneficial to control antibiotic resistance across highly prevalent bacterial species (6,7). Currently there are seven RSV vaccine formulations in Phase III clinical trials, including those targeting children, pregnant women, and the elderly (8). However, there are currently no estimates of the potential impact of RSV vaccines on preventing either antibiotic prescribing or antibiotic resistance.

In this study we evaluate the likely impact of the new generation of RSV vaccine strategies on antibiotic prescribing in England, a high income setting, and use these predictions to quantify the reduction in antibiotic resistance outcomes.

## Methods

### Literature review

We searched Pubmed using the terms: (“Respiratory Syncytial Virus, Human”[MeSH Terms] OR “Respiratory Syncytial Virus Infections”[MeSH Terms]) AND (“antibiotic” OR “antibiotics” OR “antimicrobial” OR “antimicrobials”) up to 3 November 2021. We included all those studies that evaluated the impact of an RSV vaccine on antibiotic use, or that estimated antibiotic use in the community, outpatient or long term care facility settings that was either i) coincident with respiratory infection symptoms with virologically confirmed RSV infection, or ii) attributed to RSV via statistical methods. We excluded all studies that reported antibiotic use only in hospitalised individuals or those attending emergency care, and excluded all review and commentary articles. We included patients of all ages and in any risk group.

### Vaccine strategies

We evaluated the impact of 12 potential RSV immunisation strategies on antibiotic prescribing rates in England. Specifically, we predicted the impact of a suite of vaccine strategies relying on vaccines that are currently under evaluation in clinical trials that can be administered via age groups or risk groups feasibly and affordably (9,10). These strategies are: vaccination of infants at two months (seasonally - VAC INF S, or year round - VAC INF A); vaccination during the third trimester of pregnancy (seasonally - MAT S, or year-round - MAT A); seasonal vaccination of toddlers aged 2-4 years (VAC 2-4 S), primary school children aged 5-9 years (VAC 5-9 S) or primary and secondary school children aged 5-14 years (VAC 5-14 S); and, seasonal administration of long-acting monoclonal antibodies. These monoclonal antibody strategies are, in increasing order of number of doses given: very high risk infants under 8 months currently eligible for Palivizumab (MAB VHR S) (11), high risk infants at birth as well as those currently eligible for Palivizumab (MAB HR S), high risk infants under 6 months as well as those currently eligible for Palivizumab (MAB HR S+), all infants at birth (MAB ALL S), or all infants under 6 months (MAB ALL S+).

We considered the efficacy against infection of long acting monoclonal antibodies and of maternal vaccination to be consistent with respective clinical trials, but that vaccine efficacy against infection in infants, toddlers and older children is consistent with natural infection, in the absence of clinical trial data (9) (**Table 2**). We assumed that vaccine uptake was consistent with other vaccine strategies delivered to the same age groups, and the monoclonal antibody uptake was the same as that for Palivizumab (9). We assumed that all vaccine strategies were administered in addition to Palivizumab while all monoclonal antibody strategies replaced Palivizumab.

**Table 1:** Literature review of RSV-associated antibiotic use in the community, outpatient visits, and long-term care facilities.

| Study first author (year) (reference) | Country | Population | Study Type | Reported Outcome |  |  |  |  |  |
| --- | --- | --- | --- | --- | --- | --- | --- | --- | --- |
|  |  |  |  | Rate of RSV-associated abx use in population |  | Abx use per RSV episode |  | RSV episodes per abx course |  |
|  |  |  |  | *Number of abx courses in vc RSV cases / 1000 py* | *Number of primary care abx courses attributable to RSV / 1000py* | *Number of abx courses / RSV-related primary care visit* | *% of vc RSV cases receiving abx courses* | *% of abx courses in study population with vc RSV* | *% of abx courses in background population attributable to RSV* |
| Ellis (2003) (21) | United States | Nursing home residents over 65 years presenting with fever or respiratory symptoms | Retrospective cohort | 62.4 (43.4–81.3) (Not high risk)<br>75.6 (58.7–92.5) High risk) |  |  |  | 3.3 (Not high risk)<br>2.8 (High risk) |  |
| Caram (2009) (22) | United States | LTCF for older adults presenting with RTI | Prospective cohort |  |  |  | 29% (2/7) |  |  |
| Turner (2012) (19) | Thailand (Maela) | Children (birth to 2y) in refugee population presenting with pneumonia | Prospective cohort | 240 (95% CI 220–260) |  |  |  |  |  |
| Meijboom<br>(2013)<br>(23) | Netherlands | Over 60y | Statistic<br>al<br>analysis |  |  | 0.75* |  |  |  |
| Taylor<br>(2016) (6) | United<br>Kingdom | Children<br>with<br>recorded<br>primary care<br>antibiotic<br>prescription | Statistic<br>al<br>analysis<br>of EHR |  | <6mo: 83.28<br>(55.47–102.65)<br>6–23mo: 119.16<br>(84.32–136.84)<br>2–4y: 74.95<br>(50.84–90.51)<br>5–17y: 10.91<br>(6.86–14.27) |  |  |  | <6mo: 19.7<br>6–23mo: 14.6<br>2–4y: 13.6<br>5–17y: 4.2† |
| Heikkinen<br>(2017)<br>(24) | Finland | Healthy<br>children<br>prospective<br>study<br>attending<br>outpatient<br>clinic (0–13y) | Prospect<br>ive<br>cohort |  |  |  | All: 54 (162/298)<br><1y: 91 (10/11)<br>1y: 63 (30/48)<br>2y: 64 (58/90)<br>3–6y: 48 (59/124)<br>7–13y: 20 (5/25)<br>** |  |  |
| Smithgall<br>(2020)<br>(25) | United States | Household<br>study of all<br>ages<br>presenting<br>with ARI<br>symptoms | Prospect<br>ive<br>househo<br>ld |  |  |  | 24 (16/66) |  |  |
| Toivonen<br>(2020)<br>(26) | Finland | Healthy<br>children 0–2y<br>presenting<br>with ARI | Prospect<br>ive birth<br>cohort |  |  |  | 35 (102/289) ** |  |  |
| Korsten<br>(2021)<br>(27) | Belgium/United Kingdom/Netherlands | Healthy community people 60+ years presenting with ARI | Prospective cohort |  |  |  | 2/36 (6%) |  |  |
| Fitzpatrick<br>(2021) (7) | United Kingdom | Children under 5y with recorded community antibiotic prescribing | Statistical analysis of EHR |  |  |  |  |  | All: 6.92<br>(5.59, 8.25)<br><1y: 5.16<br>(3.91, 6.41)<br>1-4y: 5.80<br>(4.55, 7.04) |

abx: antibiotic; py: person years; vc: virologically confirmed; EHR: electronic health records; ARI: acute respiratory infection
\*calculated using excess antibiotic prescriptions and GP visits from the RSV season in the Netherlands (28)
\*\*very low rate of hospitalizations so assumption that all antibiotic are community-prescribed
† from antibiotic prescriptions limited to those used against respiratory disease: broad spectrum penicillins, macrolides, tetracyclines

**Table 2:** Intervention assumptions. Each intervention was compared to status quo Palivizumab administered seasonally to very high risk infants (9).

| Intervention | Value | Notes |
| --- | --- | --- |
| *Monoclonal antibodies* |  |  |
| Delay between administration and protection (days) | None | (29) |
| Average period of protection (days) | 275 | (29) |
| Efficacy against symptomatic infection (%) | 70.1<br>(52.3–81.0) | (30) |
| Uptake (%) | 90% | Same as Palivizumab (9) |
| *Maternal vaccination\** |  |  |
| Average period of protection (days) | 133.5<br>(119.6–146.1) | Same as estimated maternal immunity |
| Efficacy against symptomatic infection (%) | 41.4<br>(4.1–64.2) | From Novavax Resvax trial (31) |
| Uptake (%) | 60 | Same as Tdap uptake in 3rd trimester (9) |
| *Childhood / adolescent vaccination* |  |  |
| Delay between administration and protection (days) | 11.4<br>(2.8–22.1) | Same as influenza antibody delay (32) |
| Average period of protection (days) | 358.9<br>(350.7–364.7) | Assumed to be same as natural immunity (9) |
| Efficacy against any infection (%) | 9-56%** | Assumed to be same as natural immunity (9) |
| Uptake (%) | 90 (<1y)<br>45 (2-4y)<br>60 (5+y) | Assumed to be same as primary series (<1y) or LAIV (2+y) (9) |

LAIV: live attenuated influenza vaccine
\* Effect on infant; effect on mother assumed the same as childhood vaccination
\*\* Mean efficacy shown for illustrative purposes. Vaccination provides temporary protection against any disease, before individual move into a reduced susceptibility state, consistent with natural exposure. The efficacy is not used explicitly in the model, but a range can be calculated as $1 - [s + (1-s)r_i]$ for $i=1,2,3$ , where $s$ is the probability of seroconversion after vaccination ( $s = 83.0\%$ (75.0–88.0%)) and $r_i$ is the risk of infection after $i$ previous exposures relative to after $i-1$ previous exposures ( $r_1 = 0.89$ (0.85–0.93), $r_2 = 0.81$ (0.74–0.85), $r_3 = 0.33$ (0.31–0.37)).

### Antibiotic courses averted

We first calculated the age-specific reduction in the number of antibiotic courses per 1000 person years due to each of these 12 vaccine strategies by multiplying the age-specific fraction of primary case visits averted with the age-specific number of primary care visits attributable to RSV that result in an antibiotic prescription.

#### i). Primary case visits averted

We calculated the average proportional reduction in primary care visits for 0-5 months, 6-23 months, 2-4 years, 5-17 years, 18+ years using a dynamic transmission model for RSV in England (9). We calculated the average reduction across a ten year time horizon after implementation of each immunisation strategy relative to the current status quo, Palivizumab administered seasonally to very high risk infants (9). For each intervention, we generated 1,000 estimates that captured uncertainty in the RSV incidence and the intervention impact via the joint posterior model parameter distribution using the efficacy and uptake parameter estimates (**Table 2**).

#### ii). RSV-attributable primary care antibiotic prescribing

In our base case analysis, we used estimates from a previous statistical attribution model that calculated the prescribing rates in primary care attributable to RSV in England and Wales (calculated as the number of antibiotic courses per 100,000 person years for the age groups 0-5mo. 6-23mo, 2-4y and 5-17y) (6). For each of the five age groups, we assumed the point value and confidence intervals to be from a triangular distribution with a mode and 95% confidence intervals, respectively and generated 1,000 samples from these distributions (**Table 3**).

**Table 3:** RSV-attributable antibiotic prescribing assumptions

|  | RSV-attributable antibiotic prescribing<br>(courses per 100,000 person years, 95% CI) |  |  |  |  |
| --- | --- | --- | --- | --- | --- |
|  | 0-5mo | 6-23mo | 2-4y | 5-17y | 18+y |
| Base case* |  |  |  |  |  |
| Analysis A | 8328<br>(5547-10265) | 11916<br>(8432-13684) | 7495<br>(5084-9051) | 1091<br>(686-1427) | 1091<br>(686-1427) |
| Sensitivity analysis** |  |  |  |  |  |
| Analysis B | 2758<br>(2086-3428) | 3350<br>(2821-3918) | 3635<br>(2876-4425) | 559<br>(306-912) | 559<br>(306-912) |
| Analysis C | 2758<br>(2086-3428) | 3350<br>(2821-3918) | 3635<br>(2876-4425) | 559<br>(306-912) | 0<br>(0-0) |

\* assumption 18+y the same as 5-17y
\*\*underlying rates <1 year (2,770, 95% CI: 2078, 3409);1-4 years (3,645 95% CI: 2,891, 4,400);
ratio of 5-17y to 2-4y same as base case values.

We performed sensitivity analyses on the RSV-attributable antibiotic prescribing. Specifically, we used an alternative study from Scotland that calculated both the total primary care prescriptions and the fraction of these prescriptions attributable to RSV for infants for 0-11mo and for 1-4yr (7) (**Table 3**). To calculate the proportion of RSV-attributable prescriptions for 5- 17y we assumed that the ratio of prescribing for those aged 5-17y to those aged 2-4y was the same as the base case study. In the first sensitivity analysis we assumed that the antibiotic prescribing rate in those aged 18+y was the same as that for those aged 5-17y. In the second sensitivity analysis we assumed that there was no RSV-attributable antibiotic prescribing for those aged 18+ (**Table 3**). We calculated the results stratified by the same age groups as our base case analysis, weighting by 2019 Scottish age group population sizes, and generated 1,000 simulations as per the base case analysis.

### Impact of averted prescribing

We first converted the averted number of antibiotic courses to averted defined daily doses (DDD) (assumed to be seven per antibiotic course (12)). We then used a previously published statistical model to calculate the population impact of the averted DDD on resistant infections, and calculated the health gain from these averted resistant infections in terms of the averted deaths, and the Disability-Adjusted Life Years gained (13,14). We then calculated the averted cost of these drug resistant infections (in 2020 GBP) by multiplying the number of averted drug resistant infections by the 2020 cost of a drug resistant infection. This cost was assumed to be $1415 in 2015 USD (15), deflated to 2014 prices, before converting to 2014 GBP using equivalent health purchasing power (16), then inflated to 2020 prices at a rate of 2.3% per year (17) giving a price of £586.83 per drug resistant infection.

### Code and analysis

We conducted our analysis using R (18), plotted our results using ggplot2, and all code is available at github.com/katiito/rsvvaccines_amr.

## Results

Our search found 285 articles, 57 of which were excluded after title and abstract screening. After full text review, ten articles met our inclusion criteria (**Table 1**). Only one of these studies evaluated the impact of RSV infection on antibiotic use in a low income setting (19), with the others conducted in the US and Europe. Studies covered a range of ages, with estimates from either children, all household members, or the elderly. Studies provided estimates of one or more of the following: the rate of RSV-associated antibiotic use in population, the rate or proportion of antibiotic use per RSV episode, and the rate or proportion of RSV episodes per antibiotic course. Studies were either retrospective or prospective cohorts or statistical attributable models. Differences in study design and reporting prevent straightforward comparisons between the estimates. For our analysis, we use the two study estimates that are both in the UK, and because of their similar study design, study population and age-stratification, lead to more comparable estimates.

We first assessed the impact of RSV immunisation strategies on primary care visits. Our model found that only four intervention strategies were able to reduce primary care visits by more than 5% in non-targeted age groups through herd protection: seasonal administration of monoclonal antibodies to <6 months (MAB ALL S+), seasonal (VAC INF S) or year-round (VAC INF A) administration of infant vaccination, and seasonal administration of vaccine to 5-14y (VAC 5-14 S). Conversely, many strategies did not reduce GP visits by 5% within the target age group (**Table 4**).

**Table 4:**
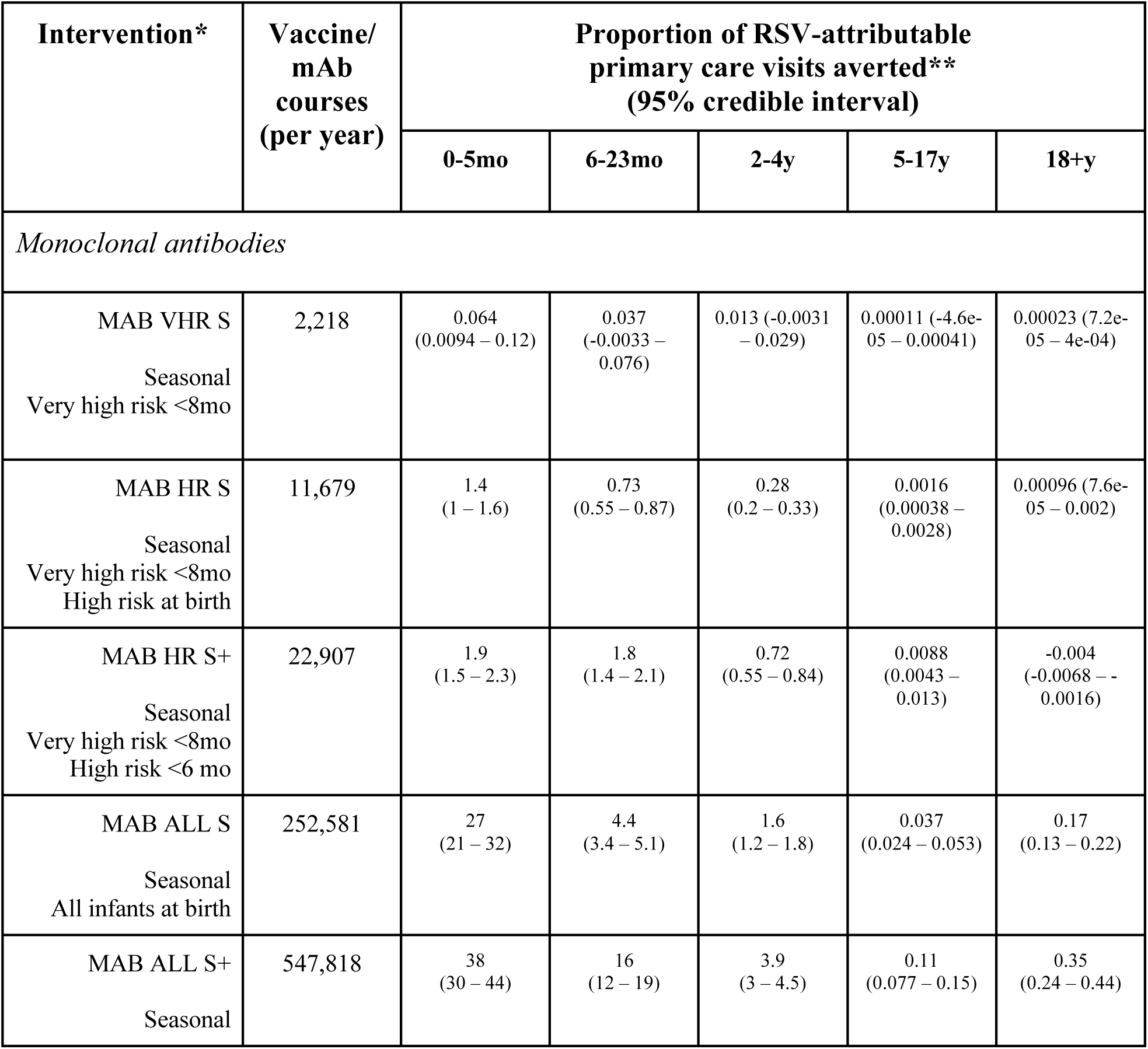
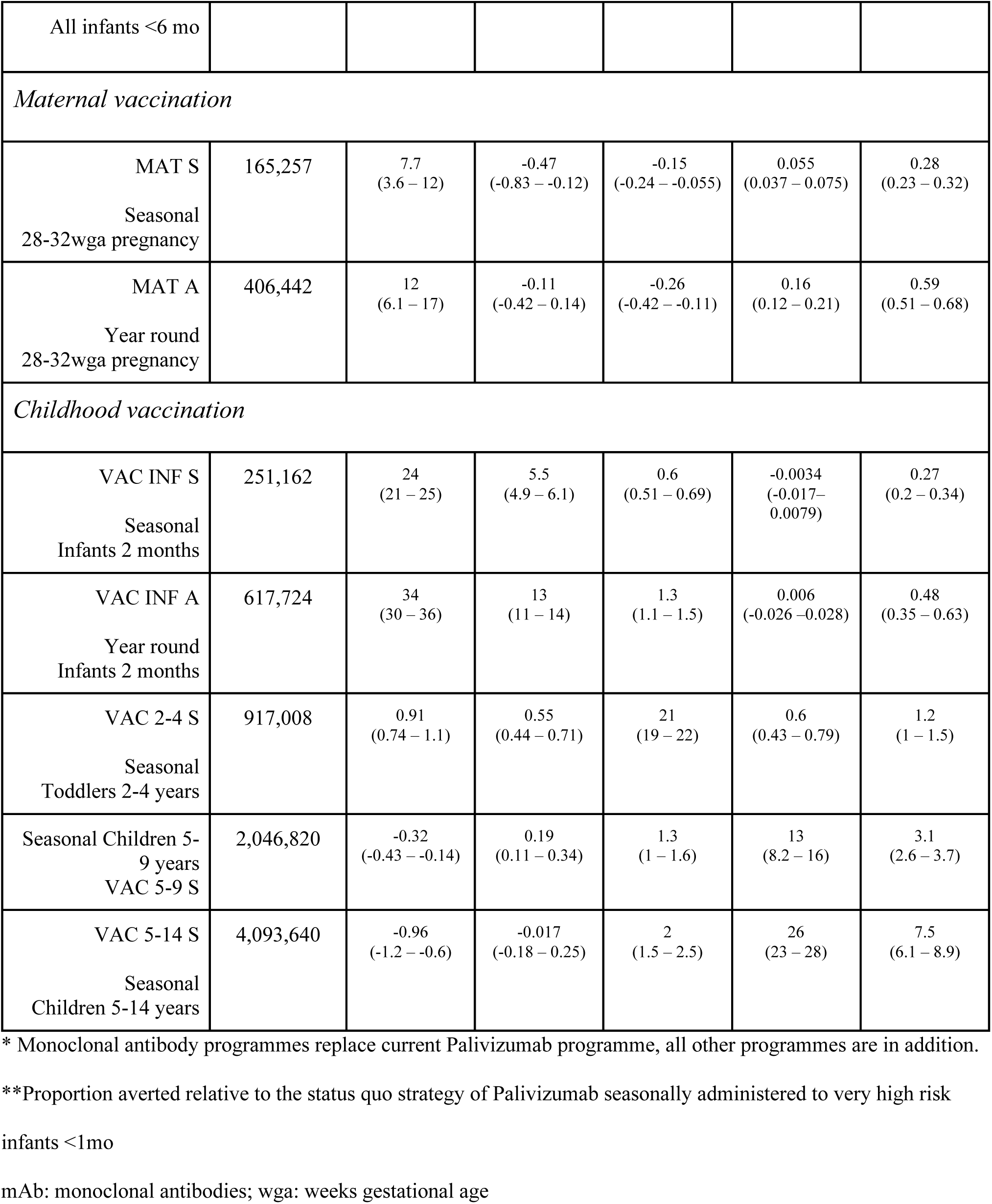
Predicted reduction in RSV-attributable primary care visits in England

There was a substantial difference in the annual number of antibiotic courses averted across both age group and intervention strategy. For infants <6 months, the largest number of courses averted were for mAb strategies administered to all infants, and for any of the infant or maternal vaccine strategies. Similarly, those aged 6-23 months benefitted from mAb or infant vaccine strategies. For older children, only vaccines that targeted their age group led to substantial benefits in annual averted antibiotic courses (**Figure 1**).

**Figure 1:**
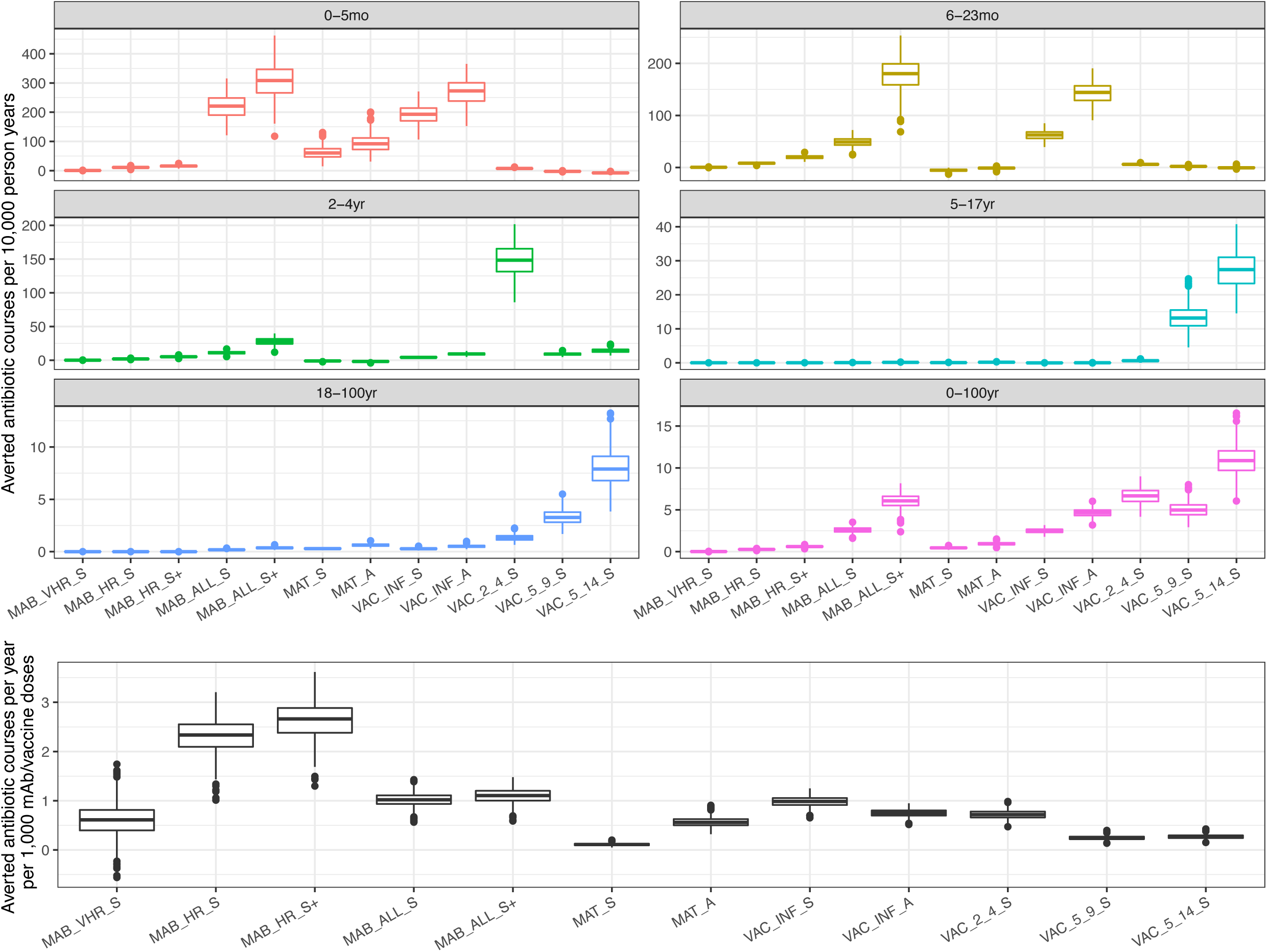
Averted antibiotic courses for each of the monoclonal antibodies (MAB), maternal vaccination (MAT) and age-targeted vaccination (VAC) interventions under base case assumptions (upper panel). Efficiency of each of the strategies at averting antibiotic courses (lower panel) under base case assumptions. Note the different y-axis scales.

The strategies that performed best at reducing antibiotic prescribing across the whole population were, in decreasing order: 5-14y vaccination (a reduction of 10.9, 95% CI: 8.0-14.2 antibiotic courses per 10,000 person years; VAC 5-14 S), 2-4y vaccination (6.6, 95% CI: 5.0-8.3; VAC 2-4 S), seasonal monoclonal antibody administration to all infants under 6 months (6.0, 95% CI:4.4-7.5; MAB ALL S+), then 5-9y vaccination (5.0, 95% CI: 3.6-6.7; VAC 5-9 S), then year-round infant vaccine (4.6, 95% CI: 3.8-5.5; VAC INF A). The remaining strategies averted on average fewer than 3 antibiotic courses per 10,000 person years across all simulations (**Figure 1**). The best performing strategy of seasonal child and adolescent vaccination (VAC 5-14 S) averted 0.23% (95% CI: 0.16-0.29%) of the total antibiotic prescriptions (for any indication or aetiology) recorded in 2018. Monoclonal antibody administration to high risk infants most efficiently reduced antibiotic use per intervention course, reducing the number of antibiotic courses per person year by 2.6 (95% CI: 1.9-3.3) per 1,000 mAb doses (**Figure 1**).

We calculated that seasonally vaccinating those aged 5-14 years (VAC 5-14 S) would lead to an annual gain of 128 (95% CI: 91-165) DALYs due to averting drug-resistant bacterial infections. This annual gain in DALYs is attributable to 86 (95% CI:61-111) infections and 4 (95% CI:3-6) deaths caused by drug-resistant bacteria (**Figure 2**). The annual averted cost of these drug resistant cases, which would be in addition to averted costs due to RSV disease, is £51,000 (95% CI:36,000-65,000) in 2020.

**Figure 2:**
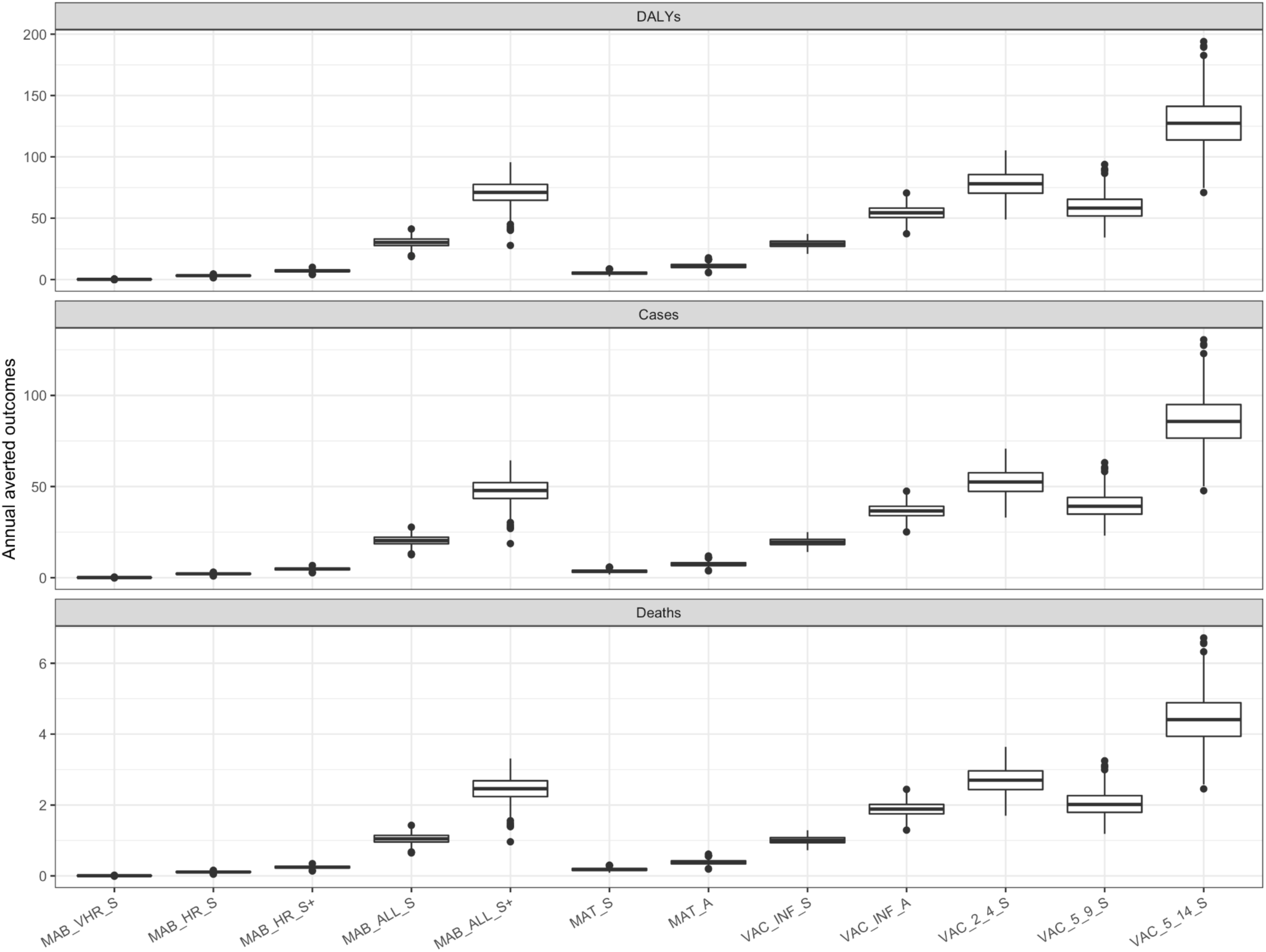
Population impact of the reduction in antibiotic prescribing for each of the monoclonal antibodies (MAB), maternal vaccination (MAT) and age-targeted vaccination (VAC) interventions under base case assumptions.

The sensitivity analyses projected less impact of RSV vaccines on antibiotic use. Under the first sensitivity analysis, the 5-14y vaccine strategy (VAC 5-14 S) averts 5.8 (95% CI: 3.1-9.5) antibiotic courses per 10,000 person years, equivalent to around 0.1% (95% CI: 0.06-0.2%) of all prescriptions in 2018 (**Figure S1, S2**). Under the alternative assumptions in the presence of no RSV-attributable prescribing over the age of 17 years, we found that toddler vaccination and child/adolescent vaccination were equally impactful in averting antibiotic courses. For comparison, under these most conservative assumptions, the child and adolescent strategy averts 2.5 (95% CI: 1.4-4.0) antibiotic courses per 10,000 person years, leading to a total annual gain of 29 DALYs (95% CI: 17-46) (**Figure S3, S4**).

## Discussion

Our study found that under the highest estimated RSV-attributable prescribing rates, the most impactful child and adolescent vaccination strategy was able to avert a quarter of a percent of the annual antibiotic prescriptions in England. This effect is limited due to a combination of factors: first, vaccination targeting the young will not substantially decrease the number of primary care visits across older age groups, so the average impact across the entire population is small. Second, antibiotic use attributable to RSV decreases with age, dropping from 20% for those under 6 months to 4% for ages 5-17 years. Finally, the assumed efficacy and uptake of the vaccine limit the total reduction in primary care visits.

Our analysis found that the combination of age-targeted RSV vaccination and age-specific variability in the RSV-associated antibiotic prescribing rates led to substantial differences in the potential reduction in antibiotic prescribing across age groups. For example, the most impactful strategy for the youngest age group, administering monoclonal antibodies to those under 6 months, would avert antibiotic prescriptions at a rate 30 times higher than is estimated in the entire population. Further, the size of the immunisation strategy largely predicts the total impact of averting antibiotic use, with the notable exception of vaccinating toddlers aged 2-4 years and monoclonal antibody administration of monoclonal antibodies to those aged less than 6 months. The impact of these two strategies across the entire population was relatively impactful due to both high coverage and efficiency.

Given increasing interest in the use of vaccines, including viral vaccines, to control antibiotic resistance, it is advisable to consider averted resistance outcomes in the economic evaluation of vaccines. However for RSV, our analysis suggests there would be a modest gain in DALYs attributable to averted drug resistant infections. Specifically, even the most impactful programme under the most optimistic scenario, a seasonal childhood vaccination for 5-14 year olds, would save £51K and 128 DALYs in drug resistant outcomes. Conversely, if this programme were to be administered at £20 per vaccine course, it would cost over £81 million and need to gain 4,100 quality-adjusted life years (QALYs) to be cost-effective in England (assuming a willingness-to-pay of £20,000/QALY). Therefore, the main benefit of RSV vaccination is likely to be in averting RSV itself, rather than averting antibiotic resistance due to by-stander selection. As noted previously (13), the analysis calculating antibiotic resistant outcomes assumes a counterfactual of no infection. Any deviation from this assumption would further reduce the impact of RSV vaccination on resistant outcomes (20).

Our results suggest that with a fixed uptake and efficacy of a vaccine, the impact of an RSV immunisation strategy on antibiotic prescribing is largely driven by the rate of antibiotic use attributable to RSV. For countries where antibiotic use due to RSV is high, the benefits of an RSV vaccine programme could be much larger and conceivably alter the cost-effectiveness of vaccination strategies. Indeed, a study in Maela, on the Thailand-Myanmar border found that young children who presented with antibiotic-indicated pneumonia who were later confirmed as RSV-infected did so at over twice the rate (240 courses given per 1000 children) (19) as children of the same age prescribed antibiotics in primary care attributable to RSV in the UK (110 courses given per 1000 children) (6). As the clinical presentation and antibiotic prescribing was not comparable between the two studies, it is likely that a vaccine targeted at children in Maela would have more than double the impact on reducing antibiotic use compared with children in the UK. Moreover, it is not only the rate of RSV-associated symptoms that determines the effect of an RSV vaccine on antibiotics, but the magnitude of total antibiotic use. For settings where over-the-counter, unregulated antibiotic use comprise the majority of drug consumption – and where, as a consequence, multidrug resistance will be higher – the benefits of RSV vaccines on controlling drug resistance will likely be substantially larger.

## Data Availability

All data produced are available online at github.com/katiito/rsvvaccines_amr

## Contributors

KEA conceived the study. KEA did the literature review and data analysis. KEA, DH, MJ, and NGD interpreted the data. DH reviewed the code. KEA wrote the manuscript, which was critically revised by DH, MJ, and NGD.

## About the authors

Dr Atkins is a Chancellor’s Fellow and Associate Professor whose work focuses on the epidemiology of antibiotic resistance and HIV.

## Transparency declarations

Nothing to declare.

## APPENDIX

### Sensitivity Analysis Results

**Figure S1:**
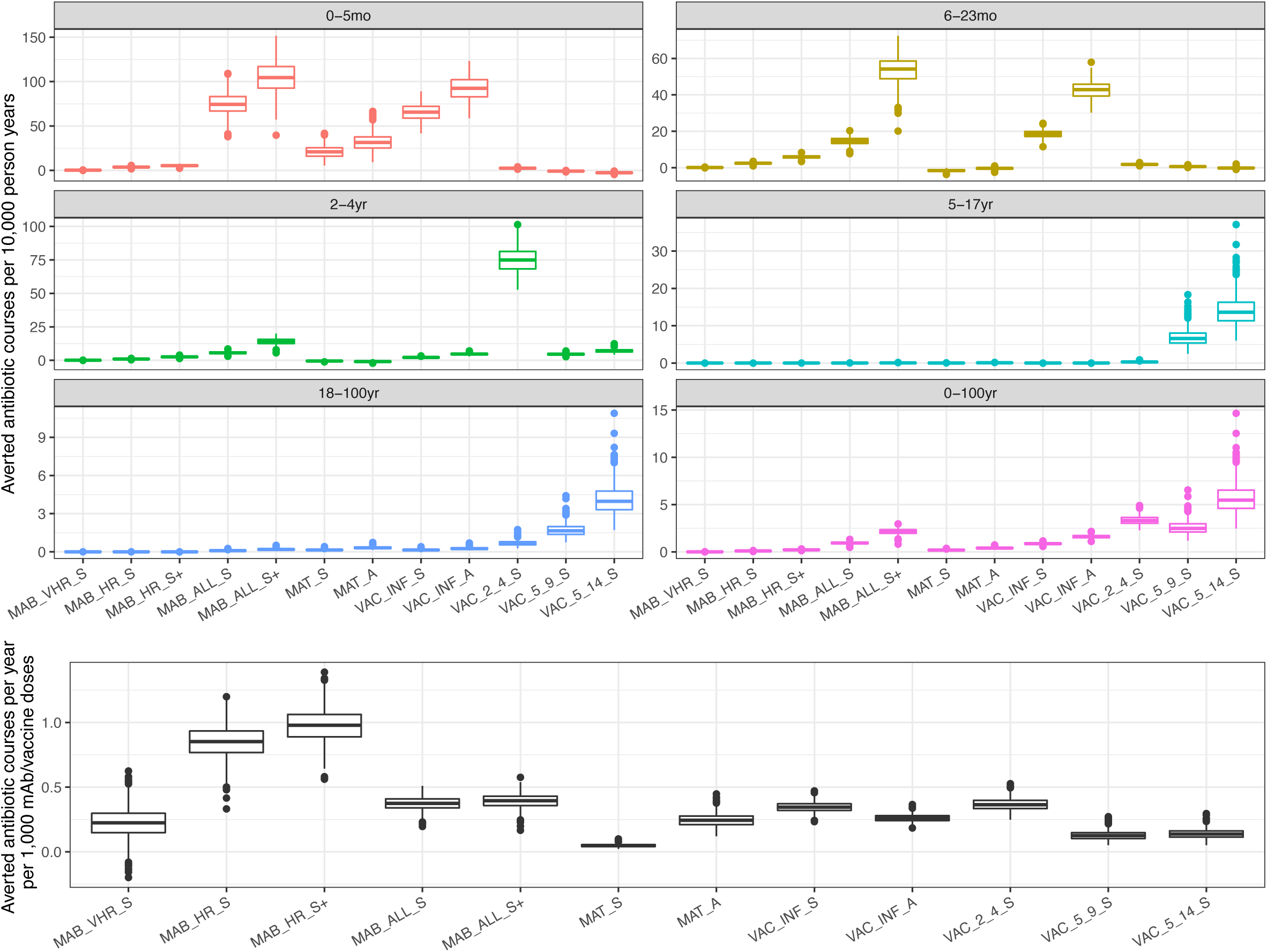
Sensitivity analysis (analysis B) of averted antibiotic courses for each of the monoclonal antibodies (MAB), maternal vaccination (MAT) and age-targeted vaccination (VAC) interventions (upper panel), and efficiency of each of the strategies at averting antibiotic courses (lower panel). Note the different y-axis scales.

**Figure S2:**
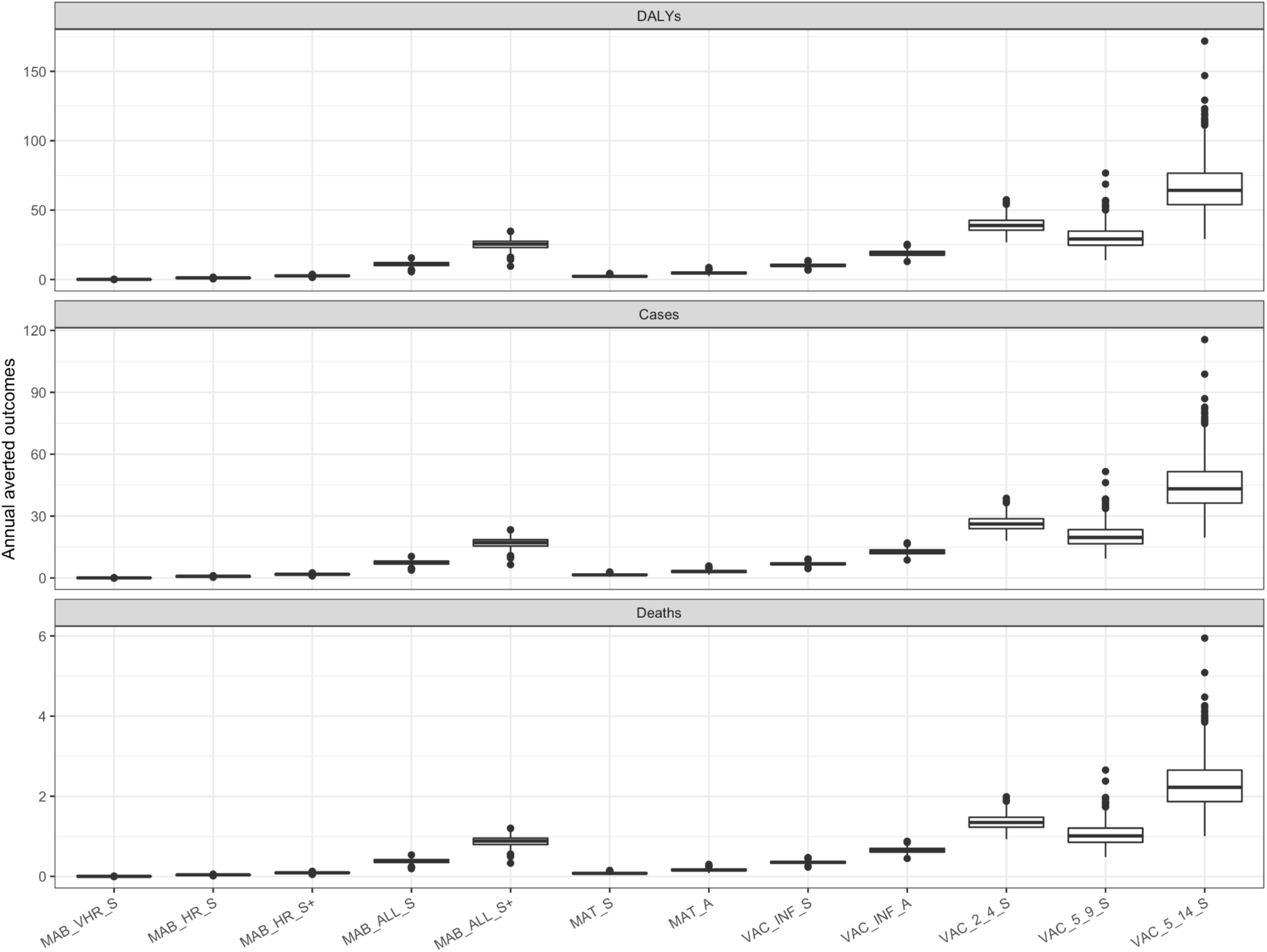
Sensitivity analysis (analysis B) for the population impact of the reduction in antibiotic prescribing for each of the monoclonal antibodies (MAB), maternal vaccination (MAT) and age-targeted vaccination (VAC) interventions.

**Figure S3:**
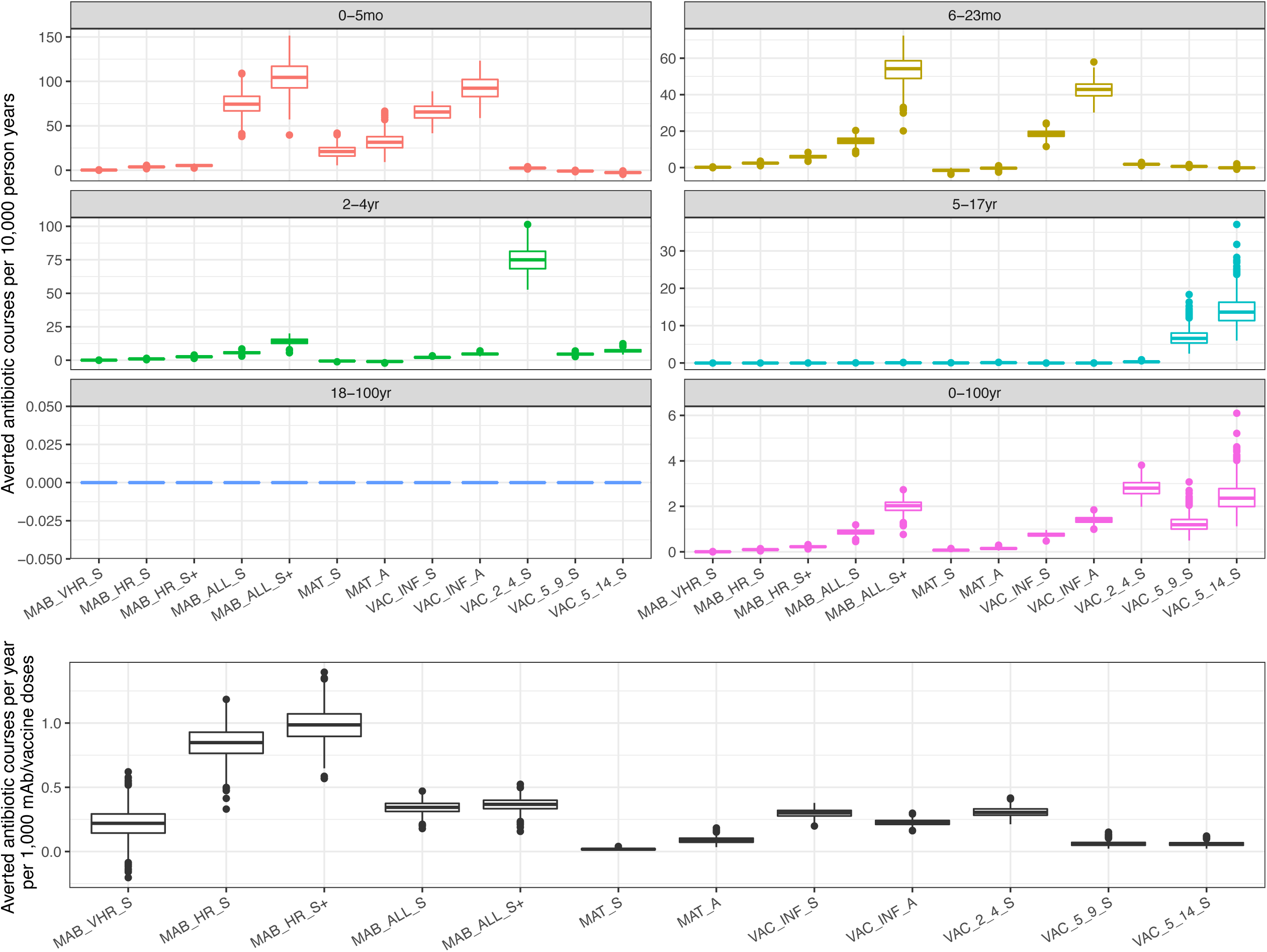
Sensitivity analysis (analysis C) of averted antibiotic courses for each of the monoclonal antibodies (MAB), maternal vaccination (MAT) and age-targeted vaccination (VAC) interventions (upper panel), and efficiency of each of the strategies at averting antibiotic courses (lower panel). Note the different y-axis scales.

**Figure S4:**
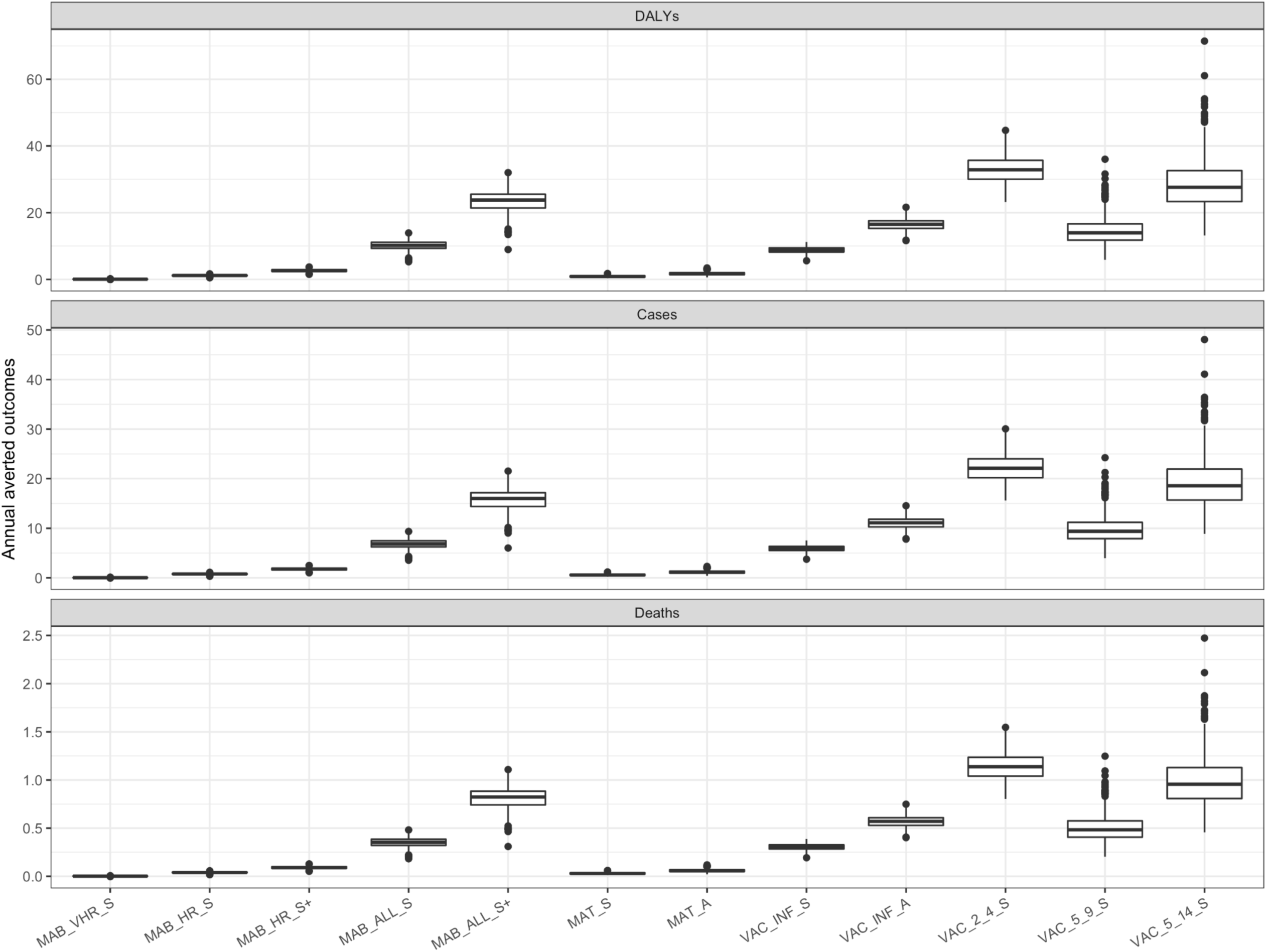
Sensitivity analysis (analysis C) for the population impact of the reduction in antibiotic prescribing for each of the monoclonal antibodies (MAB), maternal vaccination (MAT) and age-targeted vaccination (VAC) interventions.

